# Artificial intelligence in prenatal ultrasound: A systematic review of diagnostic tools for detecting congenital anomalies

**DOI:** 10.1101/2025.07.04.25330464

**Authors:** Jennifer Dunne, Chellan Kumarasamy, Daniel G Belay, Ana Pilar Betran, Amanuel T Gebremedhin, Sara Mengistu, Sylvester Dodzi Nyadanu, Aditi Roy, Gizachew A Tessema, Tamrat Tigest, Gavin Pereira

**Affiliations:** Curtin School of Population Health, Faculty of Health Sciences, Curtin University, Kent Street, Bentley, WA, 6102, Australia; Dementia Centre of Excellence, enAble Institute, Faculty of Health Sciences, Curtin University, Kent Street, Bentley WA 6102; Curtin Medical School, Faculty of Health Sciences, Curtin University, Kent Street, Bentley, WA, 6102, Australia; UNDP/UNFPA/UNICEF/WHO/World Bank Special Program of Research, Development and Research Training in Human Reproduction (HRP), Department of Sexual and Reproductive Health and Research, World Health Organization, Geneva, Switzerland; School of Nursing and Midwifery, Edith Cowan University, Perth, Australia, Curtin School of Population Health, Curtin University, Perth, Australia; School of Public Health, University of Adelaide, Adelaide, South Australia SA, 5000, Australia

**Author notes:** **Corresponding author:** Jennifer Dunne. GPO Box U1987, Perth WA 6845.

**Keywords:** congenital anomalies, pregnancy, ultrasound, artificial intelligence, machine learning

## Abstract

**Background:** Artificial intelligence (AI) has potentially shown promise in interpreting ultrasound imaging through flexible pattern recognition and algorithmic learning, but implementation in clinical practice remains limited. This study aimed to investigate the current application of AI in prenatal ultrasounds to identify congenital anomalies, and to synthesise challenges and opportunities for the advancement of AI-assisted ultrasound diagnosis. This comprehensive analysis addresses the clinical translation gap between AI performance metrics and practical implementation in prenatal care.

**Methods:** Systematic searches were conducted in eight electronic databases (CINAHL Plus, Ovid/EMBASE, Ovid/MEDLINE, ProQuest, PubMed, Scopus, Web of Science and Cochrane Library) and Google Scholar from inception to May 2025. Studies were included if they applied an AI-assisted ultrasound diagnostic tool to identify a congenital anomaly during pregnancy. This review adhered to PRISMA guidelines for systematic reviews. We evaluated study quality using the Checklist for Artificial Intelligence in Medical Imaging (CLAIM) guidelines.

**Findings:** Of 9,918 records, 224 were identified for full-text review and 20 met the inclusion criteria. The majority of studies (11/20, 55%) were conducted in China, with most published after 2020 (16/20, 80%). All AI models were developed as an assistive tool for anomaly detection or classification. Most models (85%) focused on single-organ systems: heart (35%), brain/cranial (30%), or facial features (20%), while three studies (15%) attempted multi-organ anomaly detection. Fifty percent of the included studies reported exceptionally high model performance, with both sensitivity and specificity exceeding 0.95, with AUC-ROC values ranging from 0.91 to 0.97. Most studies (75%) lacked external validation, with internal validation often limited to small training and testing datasets.

**Interpretation:** While AI applications in prenatal ultrasound showed potential, current evidence indicates significant limitations in their practical implementation. Much work is required to optimise their application, including the external validation of diagnostic models with clinical utility to have real-world implications. Future research should prioritise larger-scale multi-centre studies, developing multi-organ anomaly detection capabilities rather than the current single-organ focus, and robust evaluation of AI tools in real-world clinical settings.

## Introduction

Congenital anomalies and their associated complications account for 20-25% of perinatal deaths globally.^1, 2^ Timely and accurate diagnosis of major fetal malformations, growth disorders and placental anomalies is essential so families and health experts can make informed decisions for appropriate pregnancy management. The best non-invasive screening method for diagnosing congenital anomalies is antenatal ultrasound.^3, 4^ The World Health Organization (WHO) recommends providing one ultrasound scan before 24 weeks gestation (early ultrasound) to estimate gestational age, improve detection of fetal anomalies and multiple pregnancies, reduce induction of labour for post-term pregnancy, and improve pregnancy experience.^5^

Despite advances in technology, ultrasound has limitations in its ability to detect all fetal anomalies, with detection rates varying widely based on the severity of the anomaly and the affected organ system. The latest evidence synthesis indicates that, in low-risk or unselected women, while a first-trimester scan appears accurate in early detection of lethal and some severe fetal anomalies (91.3%), its overall anomaly detection rate is limited to 37.5%^4^. Abdominal wall anomalies had the highest detection rates: 95.6% with a first-trimester scan, 99% with combined first- and second-trimester scans, and 90.8% with a single second-trimester scan. In contrast, digestive tract anomalies had the lowest detection rates: 8.3%, 46.5%, and 33.3%, respectively.^4^ In addition, the effectiveness of fetal ultrasound heavily relies on the skill and experience of the operator and technical factors such as maternal obesity, fetal position, and equipment quality can also affect the clarity of ultrasound images.

The use of artificial intelligence (AI) in the field of radiology has considerably developed in recent years, particularly in the diagnosis of liver, thyroid, cervical and breast diseases, with increasing application in oncology-related imaging. AI is the ability of computer programs to perform processes associated with human intelligence, such as reasoning, learning, adaptation, sensory understanding and interaction.^6, 7^ Machine learning is a subset of AI that involves training models on patterns derived from data with minimal reliance on human-defined rules.^7^ A major limitation of machine learning is that it relies heavily on statistical insights and thus can be resource intensive, requiring labelling of large volumes of images to train a model. Deep learning is a branch of AI, in which convolutional neural networks (CNNs) can provide a scalable approach to image classification and object recognition,^7^ with the ability to attain a high level of performance although often requiring substantial training data. These networks process image data through multiple layers of interconnected neurons that automatically extract hierarchical features making them particularly suitable for ultrasound imaging where pattern recognition across varying tissue density is essential.

The ability of AI to interpret ultrasound imaging through flexible pattern recognition and algorithmic learning has shown great promise in the accuracy and sensitivity of image anomaly identification.^8^ Ideally suited to the medical field of radiology, the development of AI in obstetric ultrasound is currently in its infancy. Some of the unique challenges to fetal ultrasound include: the mobility of the fetus, the developing fetal anatomy, and the requirement to obtain specific planes for diagnosis which can be both difficult to obtain and limited by fetal positioning and maternal body habitus.^9^ Fetal factors such as speckle noise, occlusion of boundaries and other artifacts can also affect intelligent detection and measurement.^9^ Despite these technical obstacles, AI has the potential to automate the detection and measurement of fetal biometry and improve the accuracy of the detection of congenital anomalies. This technology could be particularly valuable in resource-limited settings where specialist expertise may be scarce, potentially bridging critical gaps in prenatal care access.

Studies leveraging AI to enhance efficiency, accuracy, accessibility and decision-making across a wide range of fields have proliferated, including those focused on prenatal diagnosis.^8–10^ While previous research has primarily focused on algorithmic development and model performance in controlled settings,^11^ this review emphasizes real-world implementation challenges. Unlike prior technical reviews, we specifically examine workflow feasibility, diagnostic utility, and equity implications of AI adoption in prenatal care. Our analysis addresses the translational gap between technical capability and clinical integration, highlighting implementation barriers in diverse healthcare systems, including low-resource contexts where ultrasound machines are not as widely accessible.^12^ This systematic review aimed to investigate the current application of AI in prenatal ultrasounds to identify congenital anomalies, and to identify challenges and opportunities for the advancement of AI-assisted ultrasound diagnosis during pregnancy for clinical utility.

## Methods

This systematic review was conducted in accordance with the Preferred Reporting for Systematic Review and Meta-Analysis (PRISMA) reporting guidelines.^13^ Additionally, we used the Checklist for Artificial Intelligence in Medical Imaging (CLAIM)^14^ to extract and evaluate key technical aspects of the AI methodologies, including model architecture, training procedures, validation approaches, and performance metrics. This dual methodological approach ensured comprehensive assessment of both the systematic review process and the specialised AI applications in prenatal ultrasound imaging. The protocol for this review was registered with PROSPERO (CRD420251012388).

### Search methods for identification of studies

A systematic search was conducted in eight electronic databases (CINAHL Plus, Ovid/EMBASE, Ovid/MEDLINE, ProQuest, PubMed, Scopus, Web of Science and Cochrane Library) from inception to 12^th^ October 2023 (Supplementary Table 1). Additionally, we searched Google Scholar for grey literature and undertook backward citation chaining of the reference list of the included studies for potentially relevant records that were not identified by the electronic search. A combination of medical subject headings, keywords and search terms were identified as relevant to AI, ultrasound diagnosis and congenital anomalies. We conducted an updated search in May 2025 using Elicit,^15^ an AI-powered research assistant based on the same inclusion and exclusion criteria.

### Eligibility criteria for inclusion

Studies were eligible for inclusion if they satisfied the following inclusion criteria: i) use of AI/machine learning/neural networks/deep learning in conjunction with ultrasound technology, ii) outcome of congenital anomalies that met the British Paediatric Association Classification of Diseases (five-digit extension of the International Classification of Diseases, Ninth Revision (ICD-9). In terms of study design, we included experimental studies (randomized controlled trials, quasi-experimental designs), observational studies (cohort, case-control, cross-sectional), and diagnostic accuracy studies that provided sufficient methodological detail to evaluate the AI application process. The study population comprised pregnant women of any risk category (low-risk, high-risk, or unselected) at any gestational age undergoing ultrasound examination. No geographic limitations were applied to ensure global representation of AI applications in prenatal diagnostics. Studies were excluded if they were: primarily focused on model development without providing any outcome assessment, published in languages other than English, not conducted in humans, and were protocols, systematic reviews, editorials, commentaries, letters or conference abstracts. Studies with insufficient methodological reporting that prevented adequate assessment of the AI implementation or validation approach were also excluded. Additionally, we excluded studies with insufficient methodological reporting that prevented adequate quality assessment of the AI implementation or validation approach, as determined by key criteria from the CLAIM checklist.

### Process for study selection and data extraction

All unique records identified from the electronic database were imported into an Endnote library and duplicate entries were removed. Covidence was used to manage the two-stage screening process. We did not use AI for screening or study selection. In the first stage of the review, two reviewers (JD & BD) screened and reviewed the titles and abstracts of all identified records retrieved during the searches and selected potentially relevant studies for full-text assessment. Three reviewers performed the full-text review of the identified articles (JD, BD, CK) such that each article was screened by two reviewers. All reviewers made decisions regarding inclusion/exclusion and the primary reason for exclusion was documented. An additional reviewer resolved any disagreements between the reviewers during each of the screening and review stages.

For each included study, data were extracted by two reviewers (JD, CK) using a data collection form which was developed by adapting common themes from other scoping reviews on AI in addition to requirements from the CLAIM^14^ (Supplementary Table 3). The data extraction tool included: author and year of publication, study aim, study design, geographic locations, participants demographics, outcome measurements (including metrics of diagnostic performance such as sensitivity, specificity, and AUC-ROC), specific types of congenital anomalies detected and sample size (distinguishing between training, validation and testing datasets), type of ultrasound and specific target (e.g. placenta), the AI model and training, methods used to evaluate the model, results, authors acknowledged limitations and implications for practice. We performed a narrative synthesis and tabular presentation of all included studies. The key characteristics of the included studies and methodology details were also tabulated and discussed. Where possible, we pooled the findings across similar congenital anomalies to present findings on the accuracy and effectiveness of the AI model on ultrasound diagnosis. When comparing AI models, we considered not only the similarity of congenital anomalies targeted but also the specific functions performed by each AI model (e.g., image quality enhancement, segmentation, classification, or diagnostic decision support) to ensure appropriate comparisons between models with similar technical objectives. A qualitative synthesis of reported implementation challenges, ethical considerations, and clinical integration factors was conducted to identify common barriers and facilitators to AI adoption in prenatal diagnostics.

## Results

Of the 9,319 records returned through bibliographic databases and an additional two studies from grey literature search, 7,143 unique records were screened for title and abstract, of which 224 were retrieved for full-text screening (see Figure 1). After full-text review, 20 studies met all inclusion criteria. The primary reasons for exclusion were studies with primary focus on model development without clinical utility (n=159), studies that did not use artificial intelligence or machine learning methods (n=37), retracted studies (n=8), and insufficient outcome reporting or inadequate methodological detail (n=20). Of the 159 studies that had a primary focus on how machine learning models were being developed to facilitate fetal ultrasounds, the most frequent application was for image segmentation, which involves identifying and highlighting specific regions or structures of interest within ultrasound images,^16^ offering no immediate diagnostic or clinical utility for sonographers.

**Figure 1.**
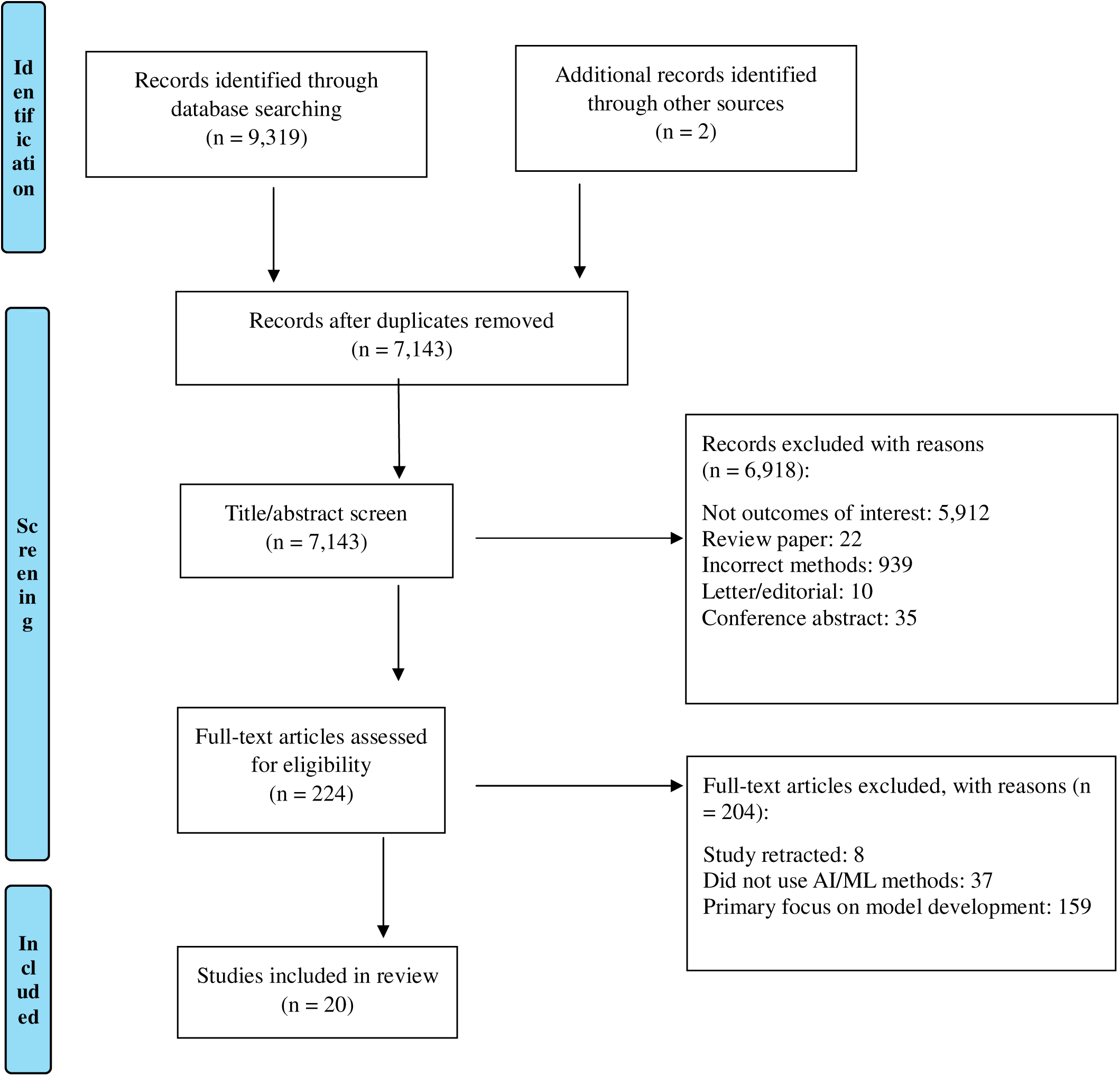
Systematic review of the literature examining the artificial intelligence diagnostic tools for detecting congenital anomalies.

### Study characteristics

Table 1 summarises the characteristics of the included studies. Among the 20 studies included, 11 (55%) were conducted in China, two (10%) in Indonesia, and the remaining studies were conducted in the USA, India, Netherlands, Tunisia, UK/France, Canada and the Czech Republic. All were conducted after 2010, with most published after 2021. The sample size varied considerably, ranging from 60 images^17^ to over 100,000 images.^18^ The fetal organ systems studied were heart (7 studies, 35%), brain/cranial (6 studies, 30%), facial features (4 studies, 20%), head and neck (2 studies, 10%), spine (1 study, 5%), and placental (1 study, 5%). Model evaluation was conducted using AUC-ROC curves in 16 (80%) studies, while accuracy was reported in 10 (50%) studies, sensitivity/specificity in 15 (75%), and F1 score in 10 (50%). Only five models (25%) were externally validated.

**Table 1.**
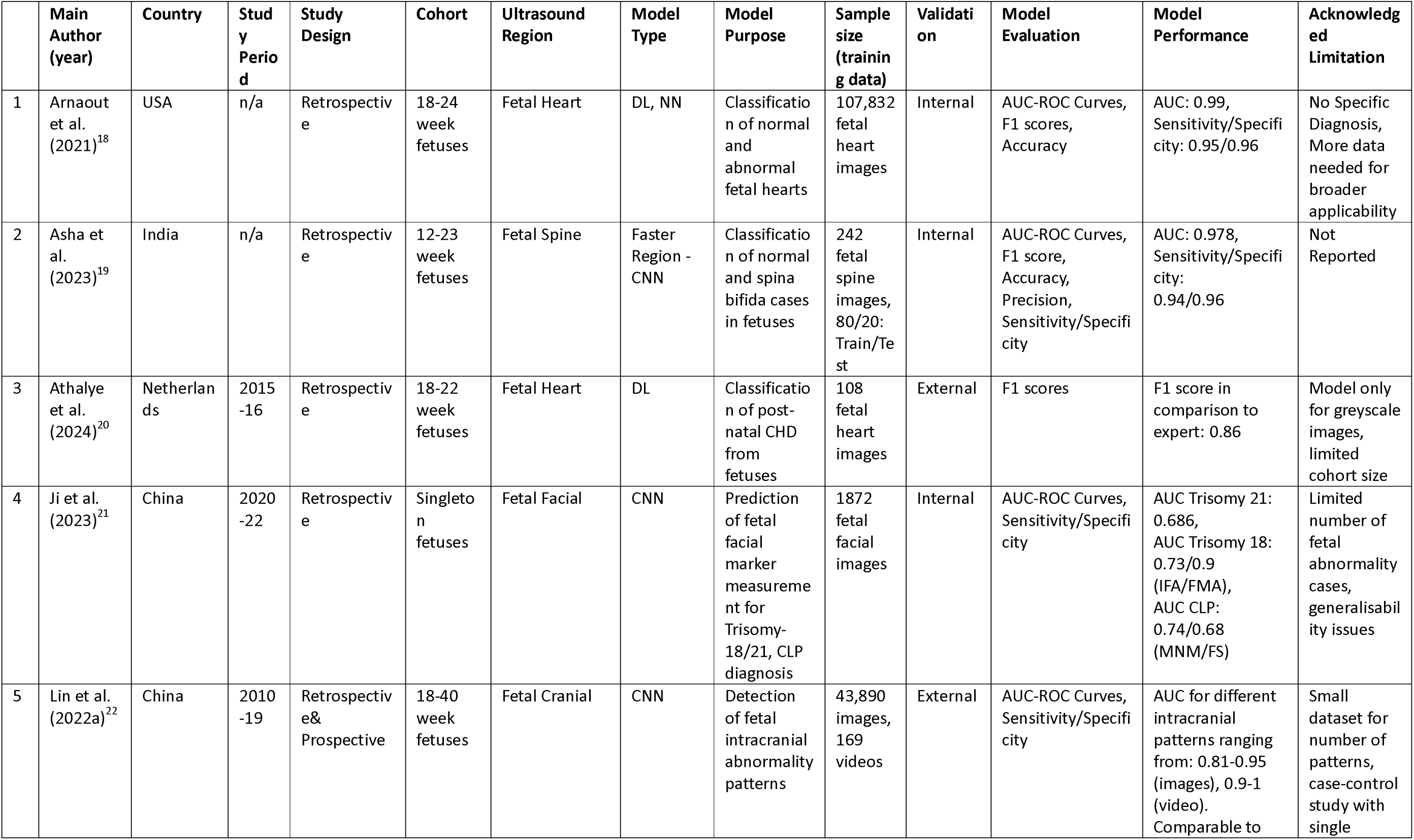
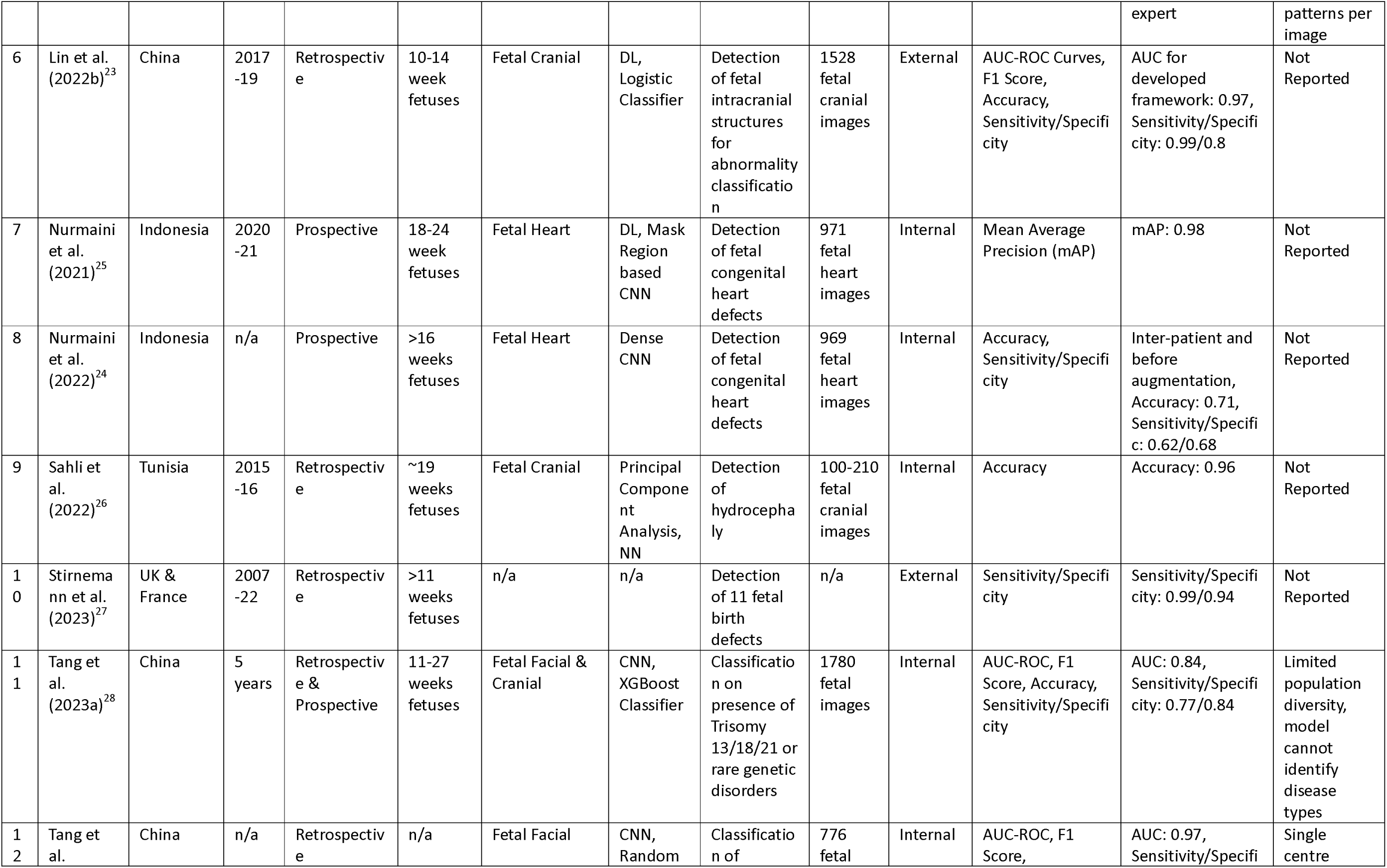
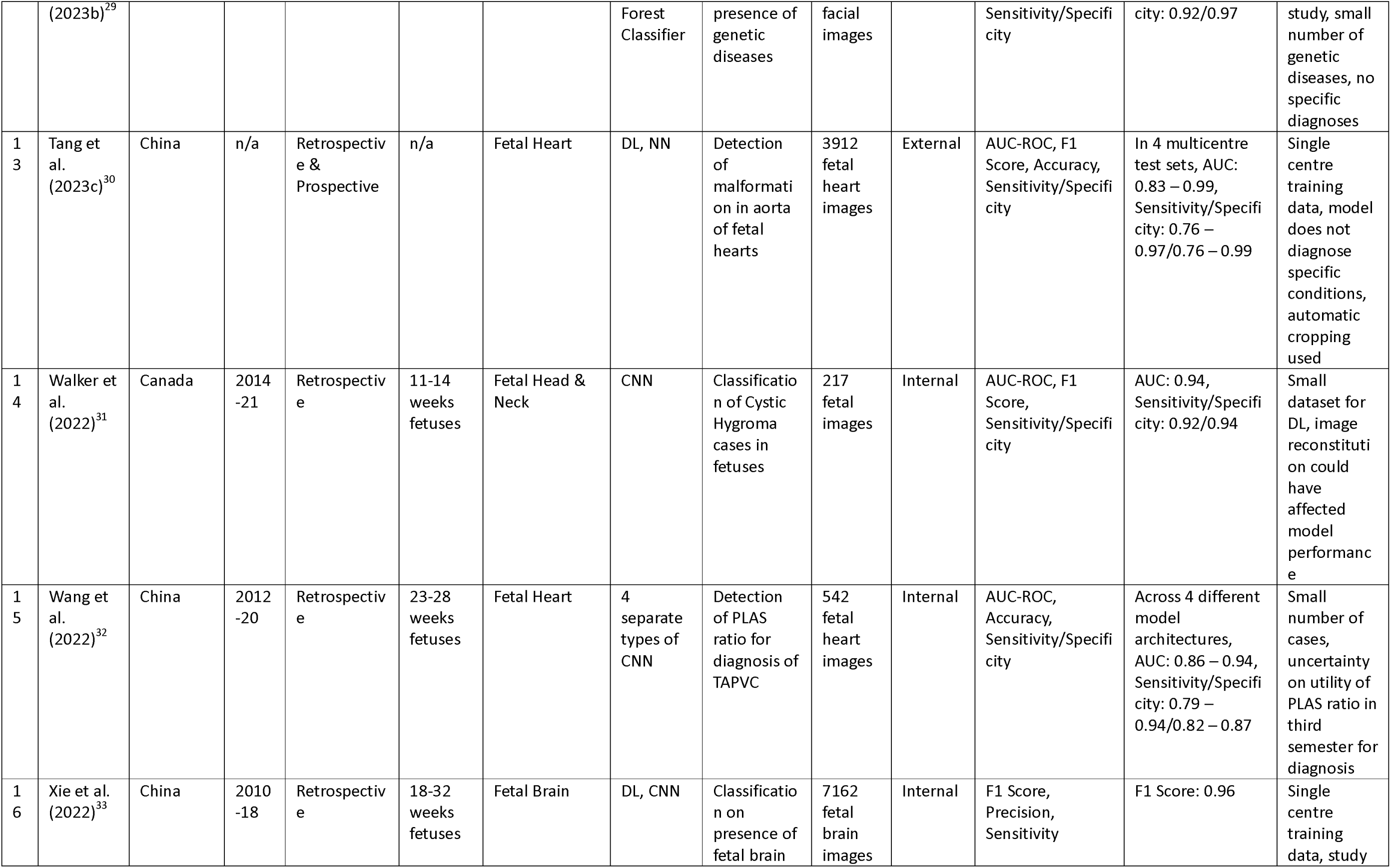
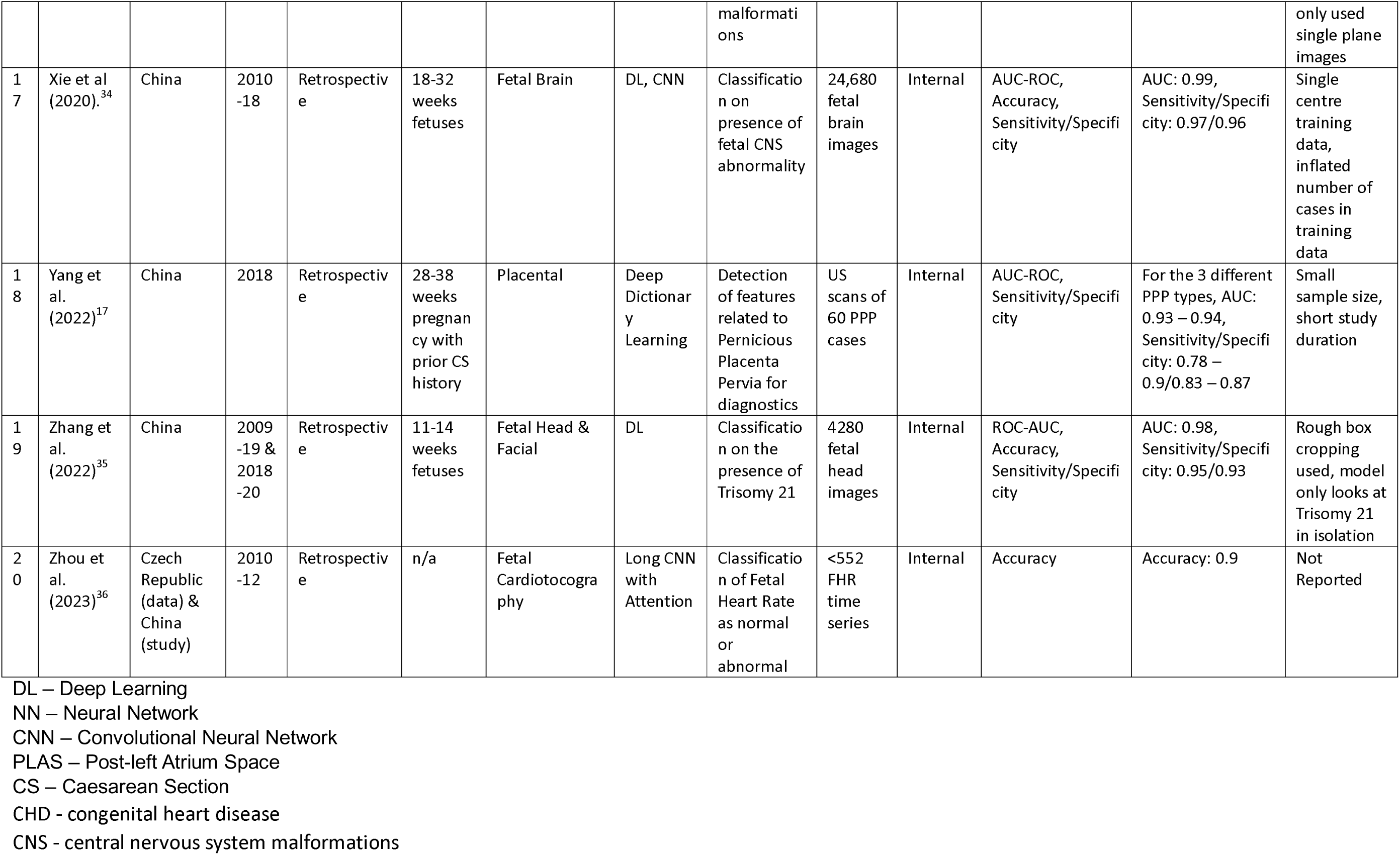
Summary of the studies applying AI in prenatal ultrasounds to identify congenital anomalies (n=20)

### AI model characteristics based on the CLAIM framework

According to the CLAIM assessment framework, we evaluated key technical aspects of the AI models across four domains: model architecture, training data, validation approaches, and clinical application.

In terms of *model architecture* and development, nearly all diagnostic models (19/20, 95%) employed image classification approaches.^17–35^ The exception was one study^36^that utilised time-series analysis of cardiotocography data. Neural networks were the predominant methodology (19/20, 95%), specifically Deep Learning Neural Networks (DL-NN) or Convolutional Neural Networks (CNN^18–25, 27–36^ Only one study^17^ diverged from this trend, employing Deep Dictionary Learning instead. Novel architectural modifications appeared in three studies: one^19^ presented a Faster Region CNN, another^24^ implemented a Mask Region CNN, and a third^36^developed a Long CNN with attention mechanism. Despite the common use of neural networks, no two studies employed identical model architectures, with considerable variation in implementation based on factors such as model purpose, dataset characteristics, and author preferences.

Regarding *training data* characteristics, the vast majority of studies (18/20, 90%) relied exclusively on still ultrasound images for model development.^17–22, 24–33, 36–38^ Only two studies explored more dynamic data sources: one^34^ incorporated ultrasound video data alongside still images for detecting intracranial abnormalities, while another^35^ employed time-series cardiotocography data to predict fetal heart abnormalities. Training dataset sizes varied substantially, ranging from just 60 images^28^ to 107,832 images,^29^ with a median size of 971 images across all studies. Most studies (15/20, 75%) used automated image segmentation algorithms or rough bounding boxes to annotate the anatomical features of interest^17, 19, 21, 24–27, 29, 31, 33–38^ with one study^30^ explicitly noting this as a methodological limitation. Only three studies^29, 32, 33^ featured expert-annotated training data by sonographers, which notably corresponded to the largest datasets in our review (ranging from 24,680 to 107,832 images).

*Validation approaches* revealed a significant limitation across the literature. Three-quarters of the studies (15/20, 75%) relied solely on internal validation^17, 19, 21, 24, 27–30, 32, 33, 35–38^ with only five studies^18, 25, 26, 31, 34^ performing external validation to verify model generalizability. All externally validated studies were conducted by research groups supported by multiple funding sources, suggesting a resource barrier to more rigorous validation. Twelve studies (60%) employed cross-validation techniques to maximize the utility of limited datasets. Only three studies (15%)^21, 24, 34^ compared AI performance against expert sonographers, providing a clinical benchmark for model evaluation.

The model purpose and *clinical application* analysis revealed significant limitations in scope and utility. Most studies (13/20, 65%) focused on binary classification (normal/abnormal) without providing specific diagnoses, while fewer (7/20, 35%) targeted detection of specific anatomical features. Notably, all studies (20/20, 100%) were limited to single-organ systems rather than providing comprehensive assessment across multiple fetal structures. Only one study^26^ attempted multi-class classification for multiple anomalies, though as this employed a commercial model, details on architecture and training were unavailable. Most studies (16/20, 80%) were restricted to gestational age ranges, further limiting clinical applicability.

### Performance metrics

Model performance was reported using various evaluation metrics across the included studies. AUC-ROC curves were the most commonly used metric (16/20 studies, 80%), followed by sensitivity/specificity (15/20, 75%), F1 scores (10/20, 50%), and accuracy (10/20, 50%). Less commonly reported metrics included precision in one study (5%)^33^ and Mean Average Precision (mAP) in one study (5%).^19^ The reported performance values were consistently high across studies. Ten studies (50%)^17, 19, 24–26, 28, 29, 32–35^ reported both sensitivity and specificity exceeding 0.95 on their respective test datasets. AUC-ROC values ranged from 0.73 to 0.99, with a median value of 0.94 across the 16 studies reporting this metric. Table 1 presents performance metrics stratified by anomaly type, with cardiac anomaly detection showing the highest median AUC (0.96), followed by neural tube defects (0.95). Five studies (25%)^17, 19, 30, 35, 38^ focused their reporting on comparing performance against alternative model architectures, primarily highlighting technical improvements achieved through novel modifications. Three studies (15%)^21, 24, 34^ evaluated their AI models against expert sonographers as a clinical benchmark, while the remaining studies relied solely on statistical performance metrics without human comparators.

Substantial methodological heterogeneity was observed across studies in terms of evaluation protocols, reference standards, and metrics reporting. This variation in methodological approaches prevented meaningful quantitative synthesis of performance across studies. We note that the high-performance metrics were predominantly reported from internal validation on test datasets that often shared similar characteristics with training data, rather than from external validation on independent datasets from different clinical environments or populations. Almost all of the included studies lacked proper external validation, with internal validation being the primary type of validation.^17–19, 21, 24, 26, 28, 29, 31–36^ The studies that internally validated their models generally used small training and testing datasets. Only five studies^20, 22, 23, 27, 30^ performed external validation in addition to internal validation. Notably, these studies were conducted by research groups supported by multiple sources of external funding.

## Discussion

### Summary of Findings

Our systematic review identified a significant gap between the technical development of AI models for prenatal ultrasound and their readiness for clinical implementation in detecting congenital anomalies. Despite extensive research activity in medical imaging AI, only 20 studies met our inclusion criteria for clinical application in prenatal anomaly detection, indicating a field still primarily focused on algorithm development rather than clinical translation. Three fundamental patterns emerged from our analysis: (i) a predominance of narrow-scope models focused on single anatomical systems rather than the comprehensive assessment needed in clinical practice; (ii) methodological limitations including homogeneous training datasets, insufficient validation across diverse populations, and reliance on still images rather than dynamic assessment; and (iii) a disproportionate emphasis on technical performance metrics with limited evaluation of real-world clinical utility. These patterns collectively indicate that while the technical foundation for AI-assisted prenatal diagnosis exists, substantial refinement is required before these tools can meaningfully support clinical decision-making. The disconnect between the high performance metrics reported and the actual readiness for clinical deployment represents the most critical finding of this review.

### Implications for clinical practice

The narrow diagnostic scope of current AI models presents a significant challenge for clinical implementation. Most of the included models focused on specific anomalies or anatomical features, ^18, 21–24, 26, 28–31^ lacking the comprehensive capabilities needed for routine prenatal screening for congenital anomalies. This limitation is particularly evident when considering complex congenital malformations that manifest in subtle ways that a narrow focused AI models may not be equipped to detect.^39^ For instance, neural tube defects, cardiac anomalies, and skeletal dysplasia often present with varying degrees of severity and can be accompanied by other structural changes.^37^ The reliance on static imaging represents a fundamental mismatch with clinical practice in prenatal diagnosis. Sonographers typically evaluate fetal structures through dynamic assessment—continuously adjusting the probe to examine anatomical structures from multiple angles and planes.^40^ While 90% of the reviewed models used only still images for training and validation, only two studies in our review utilized more dynamic data sources: one^34^ incorporated fetal ultrasound video alongside still images, while another^35^ employed time-series cardiotocography data. This static versus dynamic disparity creates a significant barrier to clinical integration, as models trained on isolated images fail to capture the temporal and spatial relationships that clinicians rely on for accurate diagnosis.

The reliability of AI performance metrics also warrants critical examination. Half of the studies in our review claimed sensitivity and specificity exceeding 0.95, suggesting near-perfect diagnostic capability. However, these results should be interpreted with considerable caution for several reasons. While AI-assisted diagnostic techniques can exhibit impressive statistics in controlled environments, these findings largely stem from retrospective analyses rather than prospective studies. This distinction is significant, as retrospective studies consistently yield higher performance metrics compared to real-time evaluations. Three key factors undermine confidence in the reported performance metrics: (i) the homogeneous nature and often small size of training datasets potentially leading to overfitting (ii), the lack of external validation in 75% of studies, and (iii) the absence of performance comparisons against expert sonographers in 85% of studies. Without prospective evaluation in diverse clinical settings with varied patient populations, there remains substantial uncertainty about how these models would perform in routine practice. The risk of diagnostic errors from overreliance on inadequately validated AI systems represents a significant patient safety concern that must be addressed before clinical implementation.

### Comparison with previous research

Our findings align with previous reviews,^3, 8, 41^ which noted significant methodological limitations in AI applications for fetal ultrasound. Specifically, similar concerns were noted in this review on the limited dataset sizes, the lack of external validation and the insufficient clarity in the reporting of model performance metrics. Another shared concern, which was evidenced in our review, was the need for clinical validation against expert sonographers, of which only 15% of our included studies compared the AI model performance to human experts. Our review extends beyond these prior findings by applying the CLAIM framework to systematically evaluate AI methodology, revealing specific gaps in model development and validation that contribute to limited clinical applicability. Through this structured assessment, we found that current AI approaches—predominantly focused on single-organ, binary classification—fundamentally misalign with clinical needs for comprehensive anomaly detection with specific diagnostic guidance. Additionally, our analysis identified implementation barriers beyond technical performance, including workflow integration challenges, the mismatch between static training data and dynamic clinical assessment, and the absence of meaningful comparison against expert clinical performance. These insights help clarify why technically impressive algorithms often fail to translate into clinically valuable tools.

### Strengths and Limitations

This review employed a comprehensive search strategy across multiple databases to identify studies applying AI techniques in prenatal ultrasound for congenital anomaly detection with precise inclusion and exclusion criteria to ensure relevance and focus to the study aim. A key strength of our review was the critical evaluation of model performance across the included studies. We systematically assessed the reported performance metrics, identifying patterns of potentially inflated results and highlighting the gap between reported performance and clinical applicability. This critical lens provides valuable context for researchers and clinicians evaluating the current state of AI applications in prenatal ultrasound, offering insights into the predominant methodologies and their relative effectiveness in this specific clinical domain. This synthesis creates a foundation for future research by identifying promising approaches and methodological gaps.

The scope of our review was constrained by the limited number of eligible studies, with only 20 studies meeting our inclusion criteria. This relatively small sample potentially limits the generalizability of our findings. Additionally, our restriction to English-language publications may have potentially excluded relevant research published in other languages; however, we were still able to analyse studies from several non-anglophone countries, including China which made up 55% of included studies. Our review did not include a quantitative synthesis or meta-analysis due to significant heterogeneity in the AI models, datasets, and performance reporting methodologies across the included studies, making direct statistical comparison challenging. The variability in study designs, model architectures, and validation approaches further restricted the comparability of results. Finally, our review primarily focused on technical and methodological aspects of AI applications, with limited exploration of implementation barriers in clinical settings. Factors such as integration with existing workflows, cost-effectiveness, and healthcare provider acceptance were not comprehensively assessed, which limits the practical applicability of our findings for clinical implementation.

### Future directions

To demonstrate genuine clinical utility, future AI models must be developed using the CLAIM checklist.^14^ The advancement of the application of AI-assisted ultrasounds requires focused attention on several key areas, including the adherence to standardised protocols for model development and validation such as CLAIM, and expansion of training datasets through multi-centre collaborations to incorporate rare and complex congenital anomalies, the integration of dynamic imaging data, creation of comprehensive diagnostic models which are externally validated, and establishment of clear clinical implementation benchmarks. A more focused approach will improve detection rates while ensuring the practical utility of AI-assisted prenatal ultrasounds. Additionally, there is a critical need to expand this research beyond high-income countries where most current studies originate. Widening the investigation to diverse global contexts would require careful consideration of how AI models reflect local datasets, population characteristics, and healthcare resource constraints to ensure equitable advances in prenatal care across different settings.

An additional consideration, and a much under-researched area, is the question of medical ethics, including the allocation of medical risk and responsibility and whether AI-assisted ultrasounds are a diagnostic tool or a decision supporting tool^40^. This is particularly complex in the context of AI-assisted prenatal ultrasound, where decisions can have profound implications for both maternal and fetal health.^42^ Key ethical considerations include determining liability when AI systems contribute to diagnostic errors, establishing clear protocols for when AI recommendations conflict with clinician judgment, and ensuring appropriate informed consent processes that explain the role of AI in diagnostic decision-making.^42^ Furthermore, ethical implementation should adhere to the World Health Organization’s six key ethical principles for AI in healthcare: protecting autonomy; promoting human well-being, safety, and public interest; ensuring transparency, explainability, and intelligibility; fostering responsibility and accountability; ensuring inclusiveness and equity; and developing AI that is responsive and sustainable.^43^

## Conclusion

While AI-assisted prenatal ultrasound shows potential to identify congenital anomalies, current evidence from this review indicates significant limitations in their practical implementation, such as a too narrow diagnostic scope, reliance on still images rather than dynamic data, lack of external validation, and insufficient diversity in training datasets. This study revealed significant gaps between the current capabilities of AI-assisted prenatal ultrasound diagnosis and the requirements for meaningful clinical implementation to identify congenital anomalies. The reported high-performance metrics of the included studies require careful interpretation given the limitations in the training datasets and validation methods. To have a real-world implication, future research must focus on conducting large-scale validation studies, developing broader anomaly detection capabilities, and rigorously evaluating AI tools under real-world clinical conditions.

## Data Availability

All data produced in the present work are contained in the manuscript

## Ethics

Due to the review nature of the present study, informed consent or approval by local ethical committees was not required.

## Acknowledgements

The authors would like to acknowledge the authors of the included primary studies.

## Contributions

JD conceived and designed the study. JD, CK, DB undertook the data analysis and verified the underlying data. JD drafted the initial version of the manuscript. All authors contributed to the interpretation of the data and critically revised the manuscript. All authors had full access to tables and figures in the study and can take responsibility for the integrity of the data and the accuracy of the data analysis. All authors read and approved the final version of the manuscript. The corresponding author attests that all listed authors meet authorship criteria and that no others meeting the criteria have been omitted.

## Data Extraction Form for Systematic Review: AI in Prenatal Ultrasound for Congenital Anomaly Detection

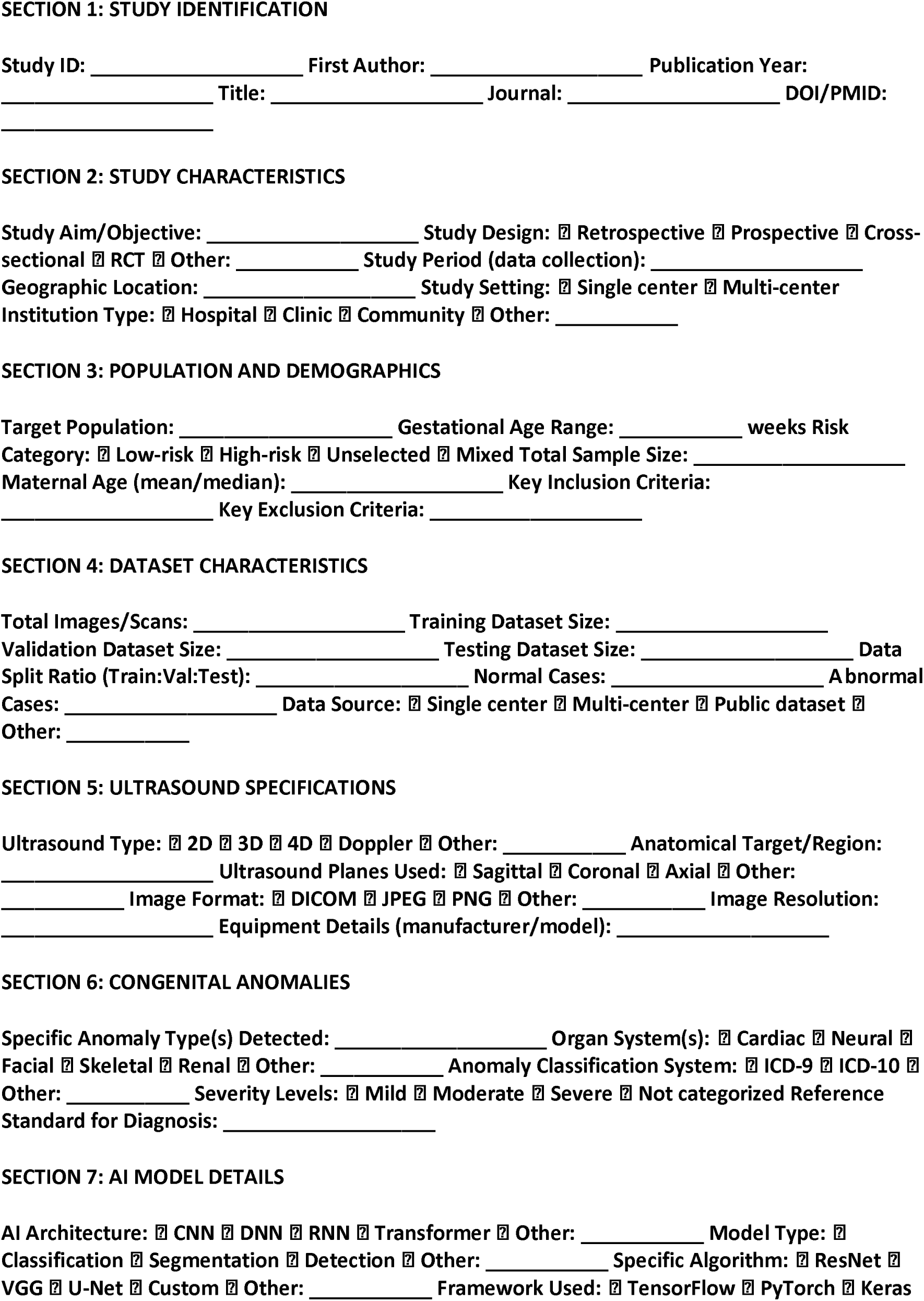
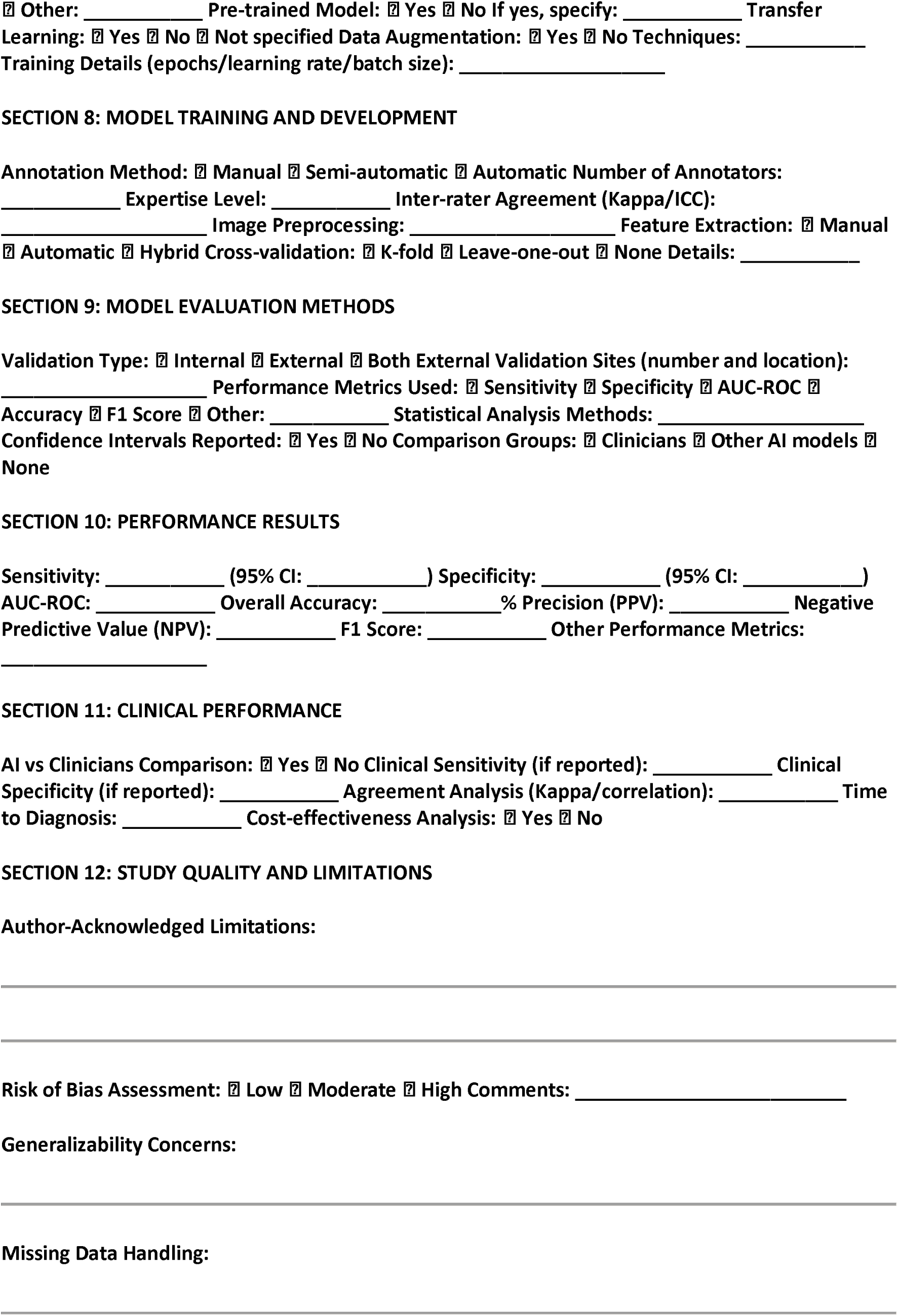
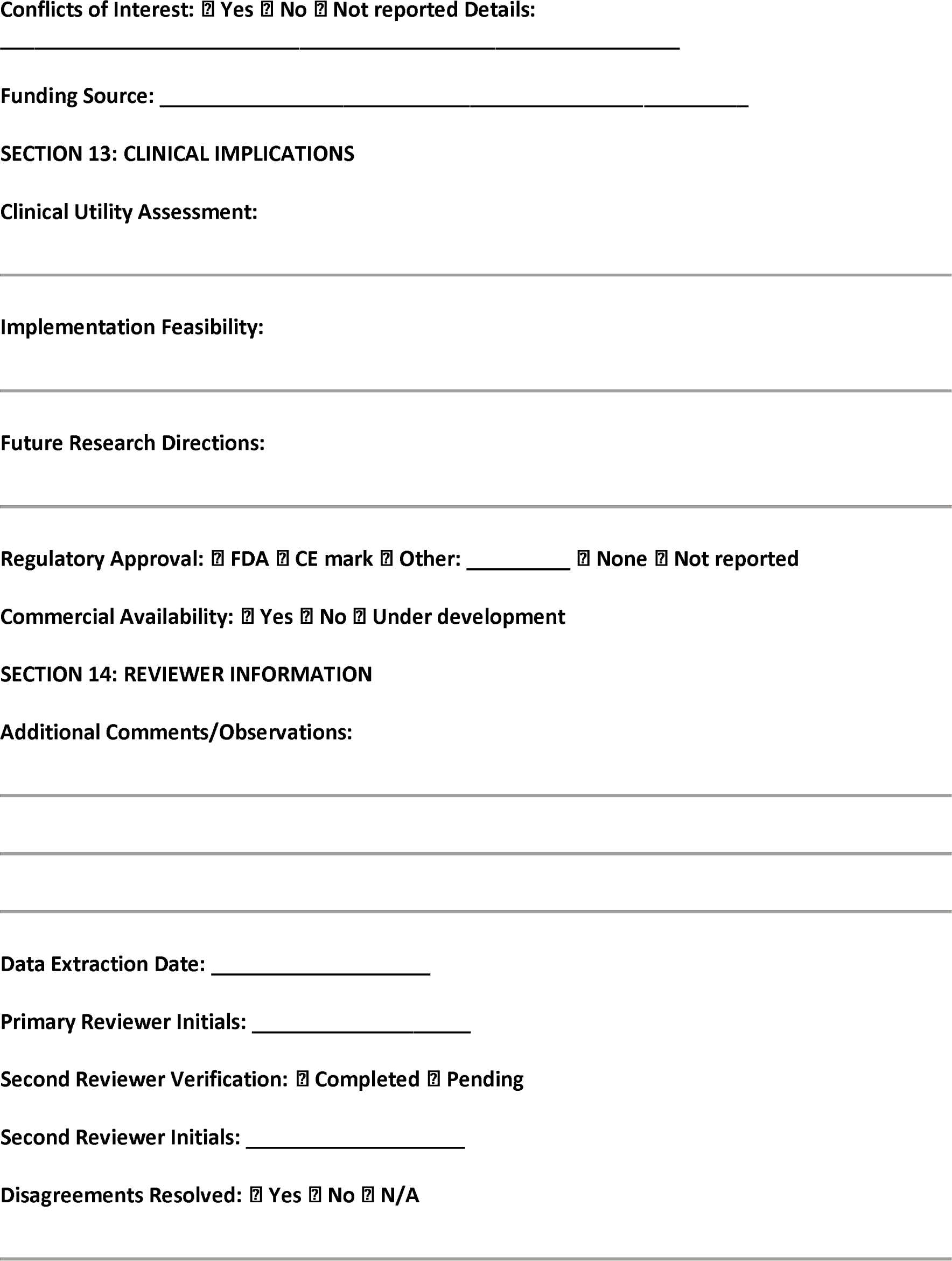

## KEY ABBREVIATIONS

AI: Artificial Intelligence
AUC-ROC: Area Under the Receiver Operating Characteristic Curve
CNN: Convolutional Neural Network
DNN: Deep Neural Network
NPV: Negative Predictive Value
PPV: Positive Predictive Value
RNN: Recurrent Neural Network
CI: Confidence Interval

