## Supplementary for "Artificial intelligence in prenatal ultrasound: A systematic review of diagnostic tools for detecting congenital anomalies"

### Supplementary Table 1. Search strategy for systematic search of the literature

| **Search engine** | **Search string** |
| --- | --- |
| CINAHL Plus | ((MH "Pregnancy+") OR (MH "Pregnancy Complications+") OR (MH "Fetal Development+") OR TI(Perinatal OR neonatal OR fetal OR foetal OR abortion OR "pregnancy termination" OR "small for gestational age" OR macrosomia OR anomalies OR malformations OR defects OR placenta* OR "intrauterine growth retardation" OR ectopic OR molar) OR AB(Perinatal OR neonatal OR fetal OR foetal OR abortion OR "pregnancy termination" OR "small for gestational age" OR macrosomia OR anomalies OR malformations OR defects OR placenta* OR "intrauterine growth retardation" OR ectopic OR molar)) AND ((MH "Artificial Intelligence+") OR (MM "Neural Networks (Computer)") OR (MH "Machine Learning+") OR TI("Artificial Intelligence" OR "Deep learning" OR "Computer-Assisted" OR "Machine learning" OR "Neural network" OR "Computational Intelligence" OR "Computer Reasoning" OR Automated OR Algorithm) OR AB("Artificial Intelligence" OR "Deep learning" OR "Computer-Assisted" OR "Machine learning" OR "Neural network" OR "Computational Intelligence" OR "Computer Reasoning" OR Automated OR Algorithm)) AND ((MH "Ultrasonography+") OR (MH "Diagnostic Imaging+") OR TI("Diagnostic Ultrasound*" OR "Ultrasound, Diagnostic" OR Diagnos* OR "Ultrasound Imaging" OR "Imaging, Ultrasound" OR Echotomography OR "Sonography, Medical" OR "Medical Sonography" OR Echography OR Echocardiography) OR AB("Diagnostic Ultrasound*" OR "Ultrasound, Diagnostic" OR Diagnos* OR "Ultrasound Imaging" OR "Imaging, Ultrasound" OR Echotomography OR "Sonography, Medical" OR "Medical Sonography" OR Echography OR Echocardiography)) |
| Ovid (Embase/Medline) | (exp pregnancy/ OR exp "pregnancy complication"/ OR exp "fetus development"/ OR (Perinatal OR neonatal OR fetal OR foetal OR abortion OR "pregnancy termination" OR "small for gestational age" OR macrosomia OR anomalies OR malformations OR defects OR placenta* OR "intrauterine growth retardation" OR ectopic OR molar).ti,ab.) AND (exp "artificial intelligence"/ OR exp "artificial neural network"/ OR exp "machine learning"/ OR ("Artificial Intelligence" OR "Deep learning" OR "Computer-Assisted" OR "Machine learning" OR "Neural network" OR "Computational Intelligence" OR "Computer Reasoning" OR Automated OR Algorithm).ti,ab.) AND (exp echography/ OR exp "diagnostic imaging"/ OR ("Diagnostic Ultrasound*" OR "Ultrasound, Diagnostic" OR Diagnos* OR "Ultrasound Imaging" OR "Imaging, Ultrasound" OR Echotomography OR "Sonography, Medical" OR "Medical Sonography" OR Echography OR Echocardiography).ti,ab.) |
| Proquest | (TITLE(perinatal OR neonatal OR fetal OR foetal OR abortion OR "pregnancy termination" OR "small for gestational age" OR macrosomia OR anomalies OR malformations OR defects OR placenta* OR "intrauterine growth retardation" OR ectopic OR molar) OR ABSTRACT(perinatal OR neonatal OR fetal OR foetal OR abortion OR "pregnancy termination" OR "small for gestational age" OR macrosomia OR anomalies OR malformations OR defects OR placenta* OR "intrauterine growth retardation" OR ectopic OR molar)) AND (TITLE("Artificial Intelligence" OR "Deep learning" OR "Computer-Assisted" OR "Machine learning" OR "Neural network" OR "Computational Intelligence" OR "Computer Reasoning" OR automated OR algorithm) OR ABSTRACT("Artificial Intelligence" OR "Deep learning" OR "Computer-Assisted" OR "Machine learning" OR "Neural network" OR "Computational Intelligence" OR "Computer Reasoning" OR automated OR algorithm)) AND (TITLE("Diagnostic Ultrasound*" OR "Ultrasound, Diagnostic" OR Diagnos* OR "Ultrasound Imaging" OR "Imaging, Ultrasound" OR echotomography OR "sonography, medical" OR "Medical Sonography" OR echography OR Echocardiography) OR ABSTRACT("Diagnostic Ultrasound*" OR "Ultrasound, Diagnostic" OR Diagnos* OR "Ultrasound Imaging" OR "Imaging, Ultrasound" OR echotomography OR "sonography, medical" OR "Medical Sonography" OR echography OR Echocardiography)) |
| PubMed | (Pregnancy[mh] OR "Pregnancy complications"[mh] OR "Fetal development"[mh] OR Perinatal[Title/Abstract] OR neonatal[Title/Abstract] OR fetal[Title/Abstract] OR foetal[Title/Abstract] OR abortion[Title/Abstract] OR "pregnancy termination"[Title/Abstract] OR "small for gestational age"[Title/Abstract] OR macrosomia[Title/Abstract] OR anomalies[Title/Abstract] OR malformations[Title/Abstract] OR defects[Title/Abstract] OR placenta*[Title/Abstract] OR "intrauterine growth retardation"[Title/Abstract] OR ectopic[Title/Abstract] OR molar[Title/Abstract]) AND ("Artificial intelligence"[mh] OR "Neural network, computer"[mh] OR "Machine learning"[mh] OR "Artificial Intelligence"[Title/Abstract] OR "Deep learning"[Title/Abstract] OR "Computer-Assisted"[Title/Abstract] OR "Machine learning"[Title/Abstract] OR "Neural network"[Title/Abstract] OR "Computational Intelligence"[Title/Abstract] OR "Computer Reasoning"[Title/Abstract] OR Automated[Title/Abstract] OR Algorithm[Title/Abstract]) AND (Ultrasonography[mh] OR "Diagnostic imaging"[mh] OR "Diagnostic Ultrasound*"[Title/Abstract] OR "Ultrasound, Diagnostic"[Title/Abstract] OR Diagnos*[Title/Abstract] OR "Ultrasound Imaging"[Title/Abstract] OR "Imaging, Ultrasound"[Title/Abstract] OR Echotomography[Title/Abstract] OR "Sonography, Medical"[Title/Abstract] OR "Medical Sonography"[Title/Abstract] OR Echography[Title/Abstract] OR Echocardiography[Title/Abstract]) |
| Scopus | TITLE-ABS-KEY(perinatal OR neonatal OR fetal OR foetal OR abortion OR "pregnancy termination" OR "small for gestational age" OR macrosomia OR anomalies OR malformations OR defects OR placenta* OR "intrauterine growth retardation" OR ectopic OR molar) AND TITLE-ABS-KEY("Artificial Intelligence" OR "Deep learning" OR "Computer-Assisted" OR "Machine learning" OR "Neural network" OR "Computational Intelligence" OR "Computer Reasoning" OR automated OR algorithm) AND TITLE-ABS-KEY("Diagnostic Ultrasound*" OR "Ultrasound, Diagnostic" OR Diagnos* OR "Ultrasound Imaging" OR "Imaging, Ultrasound" OR echotomography OR "sonography, medical" OR "Medical Sonography" OR echography OR Echocardiography) |
| Web of Science | (TI=(perinatal OR neonatal OR fetal OR foetal OR abortion OR "pregnancy termination" OR "small for gestational age" OR macrosomia OR anomalies OR malformations OR defects OR placenta* OR "intrauterine growth retardation" OR ectopic OR molar) OR AB=(perinatal OR neonatal OR fetal OR foetal OR abortion OR "pregnancy termination" OR "small for gestational age" OR macrosomia OR anomalies OR malformations OR defects OR placenta* OR "intrauterine growth retardation" OR ectopic OR molar)) AND (TI=("Artificial Intelligence" OR "Deep learning" OR "Computer-Assisted" OR "Machine learning" OR "Neural network" OR "Computational Intelligence" OR "Computer Reasoning" OR automated OR algorithm) OR AB=("Artificial Intelligence" OR "Deep learning" OR "Computer-Assisted" OR "Machine learning" OR "Neural network" OR "Computational Intelligence" OR "Computer Reasoning" OR automated OR algorithm)) AND (TI=("Diagnostic Ultrasound*" OR "Ultrasound, Diagnostic" OR Diagnos* OR "Ultrasound Imaging" OR "Imaging, Ultrasound" OR echotomography OR "sonography, medical" OR "Medical Sonography" OR echography OR Echocardiography) OR AB=("Diagnostic Ultrasound*" OR "Ultrasound, Diagnostic" OR Diagnos* OR "Ultrasound Imaging" OR "Imaging, Ultrasound" OR echotomography OR "sonography, medical" OR "Medical Sonography" OR echography OR Echocardiography)) |
| Cochrane Library | ([Pregnancy] explode all trees OR [Pregnancy Complications] explode all trees OR [Fetal Development] explode all trees OR (perinatal OR neonatal OR fetal OR foetal OR abortion OR "pregnancy termination" OR "small for gestational age" OR macrosomia OR anomalies OR malformations OR defects OR placenta* OR "intrauterine growth retardation" OR ectopic OR molar):ti,ab,kw) AND ([Artificial Intelligence] explode all trees OR [Neural Networks, Computer] explode all trees OR [Machine Learning] explode all trees OR ("Artificial Intelligence" OR "Deep learning" OR "Computer-Assisted" OR "Machine learning" OR "Neural network" OR "Computational Intelligence" OR "Computer Reasoning" OR automated OR algorithm):ti,ab,kw) AND ([Ultrasonography] explode all trees OR [Diagnostic Imaging] explode all trees OR ((Diagnostic NEXT Ultrasound*) OR "Ultrasound, Diagnostic" OR Diagnos* OR "Ultrasound Imaging" OR "Imaging, Ultrasound" OR echotomography OR "sonography, medical" OR "Medical Sonography" OR echography OR echocardiography):ti,ab,kw) |
| Google Scholar | ("perinatal" OR "neonatal" OR "fetal" OR "pregnancy") AND ("artificial intelligence" OR "machine learning" OR "deep learning") AND ("ultrasound" OR "diagnostic imaging" OR "sonography") |

### **Supplementary Table 2.** Preferred Reporting for Systematic Review and Meta-Analysis (PRISMA) 2020 completed checklist

| **Section and Topic** | **Item #** | **Checklist item** | **Location where item is reported** |
| --- | --- | --- | --- |
| **TITLE** | | |  |
| Title | 1 | Identify the report as a systematic review. | Pg 1, line 1 |
| **ABSTRACT** | | |  |
| Abstract | 2 | See the PRISMA 2020 for Abstracts checklist. | Pg 2, line 1 |
| **INTRODUCTION** | | |  |
| Rationale | 3 | Describe the rationale for the review in the context of existing knowledge. | Pg 4, line 1 |
| Objectives | 4 | Provide an explicit statement of the objective(s) or question(s) the review addresses. | Pg 4, line 39 |
| **METHODS** | | |  |
| Eligibility criteria | 5 | Specify the inclusion and exclusion criteria for the review and how studies were grouped for the syntheses. | Pg 5, line 17 |
| Information sources | 6 | Specify all databases, registers, websites, organisations, reference lists and other sources searched or consulted to identify studies. Specify the date when each source was last searched or consulted. | Pg 5, line 1 |
| Search strategy | 7 | Present the full search strategies for all databases, registers and websites, including any filters and limits used. | Supp T S1 |
| Selection process | 8 | Specify the methods used to decide whether a study met the inclusion criteria of the review, including how many reviewers screened each record and each report retrieved, whether they worked independently, and if applicable, details of automation tools used in the process. | Pg 5, line 28 |
| Data collection process | 9 | Specify the methods used to collect data from reports, including how many reviewers collected data from each report, whether they worked independently, any processes for obtaining or confirming data from study investigators, and if applicable, details of automation tools used in the process. | Pg 5, line 29 |
| Data items | 10a | List and define all outcomes for which data were sought. Specify whether all results that were compatible with each outcome domain in each study were sought (e.g. for all measures, time points, analyses), and if not, the methods used to decide which results to collect. | Pg 5, line 20 |
|  | 10b | List and define all other variables for which data were sought (e.g. participant and intervention characteristics, funding sources). Describe any assumptions made about any missing or unclear information. | Pg 5, line 38 |
| Study risk of bias assessment | 11 | Specify the methods used to assess risk of bias in the included studies, including details of the tool(s) used, how many reviewers assessed each study and whether they worked independently, and if applicable, details of automation tools used in the process. | Pg 5, line 43 |
| Effect measures | 12 | Specify for each outcome the effect measure(s) (e.g. risk ratio, mean difference) used in the synthesis or presentation of results. | NA |
| Synthesis methods | 13a | Describe the processes used to decide which studies were eligible for each synthesis (e.g. tabulating the study intervention characteristics and comparing against the planned groups for each synthesis (item #5)). | Pg 6, line 11 |
|  | 13b | Describe any methods required to prepare the data for presentation or synthesis, such as handling of missing summary statistics, or data conversions. | NA |
|  | 13c | Describe any methods used to tabulate or visually display results of individual studies and syntheses. | Pg 6, line 10 |
|  | 13d | Describe any methods used to synthesize results and provide a rationale for the choice(s). If meta-analysis was performed, describe the model(s), method(s) to identify the presence and extent of statistical heterogeneity, and software package(s) used. | Pg 6, line 17 |
|  | 13e | Describe any methods used to explore possible causes of heterogeneity among study results (e.g. subgroup analysis, meta-regression). | NA |
|  | 13f | Describe any sensitivity analyses conducted to assess robustness of the synthesized results. | NA |
| Reporting bias assessment | 14 | Describe any methods used to assess risk of bias due to missing results in a synthesis (arising from reporting biases). | NA |
| Certainty assessment | 15 | Describe any methods used to assess certainty (or confidence) in the body of evidence for an outcome. | NA |
| **RESULTS** | | |  |
| Study selection | 16a | Describe the results of the search and selection process, from the number of records identified in the search to the number of studies included in the review, ideally using a flow diagram. | Figure 1 |
|  | 16b | Cite studies that might appear to meet the inclusion criteria, but which were excluded, and explain why they were excluded. | NA |
| Study characteristics | 17 | Cite each included study and present its characteristics. | Supp T S1 |
| Risk of bias in studies | 18 | Present assessments of risk of bias for each included study. | NA |
| Results of individual studies | 19 | For all outcomes, present, for each study: (a) summary statistics for each group (where appropriate) and (b) an effect estimate and its precision (e.g. confidence/credible interval), ideally using structured tables or plots. | Table 1 |
| Results of syntheses | 20a | For each synthesis, briefly summarise the characteristics and risk of bias among contributing studies. | Table 1  Pg 5, line 2, line 8 |
|  | 20b | Present results of all statistical syntheses conducted. If meta-analysis was done, present for each the summary estimate and its precision (e.g. confidence/credible interval) and measures of statistical heterogeneity. If comparing groups, describe the direction of the effect. | NA |
|  | 20c | Present results of all investigations of possible causes of heterogeneity among study results. | NA |
|  | 20d | Present results of all sensitivity analyses conducted to assess the robustness of the synthesized results. | NA |
| Reporting biases | 21 | Present assessments of risk of bias due to missing results (arising from reporting biases) for each synthesis assessed. | Pg 5, line 8 |
| Certainty of evidence | 22 | Present assessments of certainty (or confidence) in the body of evidence for each outcome assessed. | NA |
| **DISCUSSION** | | |  |
| Discussion | 23a | Provide a general interpretation of the results in the context of other evidence. | Pg 13, line 5 |
|  | 23b | Discuss any limitations of the evidence included in the review. | Pg 10, line 1 |
|  | 23c | Discuss any limitations of the review processes used. | Pg 10, line 2 |
|  | 23d | Discuss implications of the results for practice, policy, and future research. | Pg 9, line 48 |
| **OTHER INFORMATION** | | |  |
| Registration and protocol | 24a | Provide registration information for the review, including register name and registration number, or state that the review was not registered. | Pg 3, line 44 |
|  | 24b | Indicate where the review protocol can be accessed, or state that a protocol was not prepared. | Pg 3, line 45 |
|  | 24c | Describe and explain any amendments to information provided at registration or in the protocol. | Pg 5, line 6 |
| Support | 25 | Describe sources of financial or non-financial support for the review, and the role of the funders or sponsors in the review. | Pg 14, line 9 |
| Competing interests | 26 | Declare any competing interests of review authors. | Pg 14, line 7 |
| Availability of data, code and other materials | 27 | Report which of the following are publicly available and where they can be found: template data collection forms; data extracted from included studies; data used for all analyses; analytic code; any other materials used in the review. | NA |

*From:* Page MJ, McKenzie JE, Bossuyt PM, Boutron I, Hoffmann TC, Mulrow CD, et al. The PRISMA 2020 statement: an updated guideline for reporting systematic reviews. BMJ 2021;372:n71. doi: 10.1136/bmj.n71

### **Supplementary Table 3.** Checklist for Artificial Intelligence in Medical Imaging (CLAIM): 2024 Update

| Section / Topic | No. | Item | Page / Line | No | NA |
| --- | --- | --- | --- | --- | --- |
| TITLE / ABSTRACT |  |  |  |  |  |
|  | **1** | Identification as a study of AI methodology, specifying the category of technology used (e.g., deep learning) |  |  |  |
| ABSTRACT |  |  |  |  |  |
|  | **2** | Summary of study design, methods, results, and conclusions |  |  |  |
| INTRODUCTION |  |  |  |  |  |
|  | **3** | Scientific and/or clinical background, including the intended use and role of the AI approach |  |  |  |
|  | **4** | Study aims, objectives, and hypotheses |  |  |  |
| METHODS |  |  |  |  |  |
| *Study Design* | **5** | Prospective or retrospective study |  |  |  |
|  | **6** | Study goal |  |  |  |
| *Data* | **7** | Data sources |  |  |  |
|  | **8** | Inclusion and exclusion criteria |  |  |  |
|  | **9** | Data pre-processing |  |  |  |
|  | **10** | Selection of data subsets |  |  |  |
|  | **11** | De-identification methods |  |  |  |
|  | **12** | How missing data were handled |  |  |  |
|  | **13** | Image acquisition protocol |  |  |  |
| *Reference Standard* | **14** | Definition of method(s) used to obtain reference standard |  |  |  |
|  | **15** | Rationale for choosing the reference standard |  |  |  |
|  | **16** | Source of reference standard annotations |  |  |  |
|  | **17** | Annotation of test set |  |  |  |
|  | **18** | Measures of inter- and intra-rater variability of features described by the annotators |  |  |  |
| *Data Partitions* | **19** | How data were assigned to partitions |  |  |  |
|  | **20** | Level at which partitions are disjoint |  |  |  |
| *Testing Data* | **21** | Intended sample size |  |  |  |

| Section / Topic | No. | Item | Page / Line | No | NA |
| --- | --- | --- | --- | --- | --- |
| *Model* | **22** | Detailed description of model |  |  |  |
|  | **23** | Software libraries, frameworks, and packages |  |  |  |
|  | **24** | Initialization of model parameters |  |  |  |
| *Training* | **25** | Details of training approach |  |  |  |
|  | **26** | Method of selecting the final model |  |  |  |
|  | **27** | Ensembling techniques |  |  |  |
| *Evaluation* | **28** | Metrics of model performance |  |  |  |
|  | **29** | Statistical measures of significance and uncertainty |  |  |  |
|  | **30** | Robustness or sensitivity analysis |  |  |  |
|  | **31** | Methods for explainability or interpretability |  |  |  |
|  | **32** | Evaluation on internal data |  |  |  |
|  | **33** | Testing on external data |  |  |  |
|  | **34** | Clinical trial registration |  |  |  |
| RESULTS |  |  |  |  |  |
| *Data* | **35** | Numbers of patients or examinations included and excluded |  |  |  |
|  | **36** | Demographic and clinical characteristics of cases in each partition |  |  |  |
| *Model performance* | **37** | Performance metrics and measures of statistical uncertainty |  |  |  |
|  | **38** | Estimates of diagnostic performance and their precision |  |  |  |
|  | **39** | Failure analysis of incorrect results |  |  |  |
| DISCUSSION |  |  |  |  |  |
|  | **40** | Study limitations |  |  |  |
|  | **41** | Implications for practice, including intended use and/or clinical role |  |  |  |
| OTHER INFORMATION |  |  |  |  |  |
|  | **42** | Provide a reference to the full study protocol or to additional technical details |  |  |  |
|  | **43** | Statement about the availability of software, trained model, and/or data |  |  |  |
|  | **44** | Sources of funding and other support; role of funders |  |  |  |

* Indicate page and/or line number for each checklist item that is present. NA = not applicable.
